# Evaluating the role of common risk variation in the recurrence risk of schizophrenia in multiplex schizophrenia families

**DOI:** 10.1101/2021.06.21.21259285

**Authors:** Mohammad Ahangari, Amanda E. Gentry, Irish Schizophrenia Genomics Consortium, Tan-Hoang Nguyen, Robert Kirkpatrick, Brian C. Verrelli, Silviu-Alin Bacanu, Kenneth S. Kendler, Bradley T. Webb, Brien P. Riley

## Abstract

Multiplex families have higher recurrence risk of schizophrenia compared to the families of sporadic cases, but the source of this increased recurrence risk is unknown. We used schizophrenia genome-wide association study data (N=156,509) to construct polygenic risk scores (PRS) in 1,005 individuals from 257 multiplex schizophrenia families, 2,114 ancestry-matched sporadic cases, and 2,205 population controls, to evaluate whether increased PRS can explain the higher recurrence risk of schizophrenia in multiplex families compared to ancestry-matched sporadic cases. Using mixed-effects logistic regression with family structure modeled as a random effect, we show that SCZ PRS in familial cases does not differ significantly from sporadic cases either with, or without family history (FH) of psychotic disorders (All sporadic cases *p* = 0.92, FH+ cases *p* = 0.88, FH-cases *p* = 0.82). These results indicate that increased burden of common schizophrenia risk variation as indexed by current SCZ PRS, is unlikely to account for the higher recurrence risk of schizophrenia in multiplex families. In the absence of elevated PRS, segregation of rare risk variation or environmental influences unique to the families may explain the increased familial recurrence risk. These findings also further validate a genetically influenced psychosis spectrum, as shown by a continuous increase of common SCZ risk variation burden from unaffected relatives to schizophrenia cases in multiplex families. Finally, these results suggest that common risk variation loading are unlikely to be predictive of schizophrenia recurrence risk in the families of index probands, and additional components of genetic risk must be identified and included in order to improve recurrence risk prediction.

## Introduction

Schizophrenia (SCZ) is a severe, clinically heterogeneous psychiatric disorder with a population prevalence of ∼1% (1). Twin, family, and adoption studies consistently show a strong genetic component, with heritability estimates of around 0.75-0.80 (2–6), and family history (FH) remains the strongest risk factor for developing SCZ (7). Despite high heritability, ∼2/3 of SCZ cases report no FH of psychotic illness, and most subjects with a positive FH (FH+) report only a single affected relative (8,9), concordant with the rates of 31% FH+ and 69% family history negative (FH-) observed in the sample of sporadic SCZ cases analyzed in this study (10).

The Irish Study of High-Density Schizophrenia Families sample (ISHDSF) (11–14) consists of 257 multiplex SCZ families with genotype data, ascertained to have two or more first-degree relatives meeting the Diagnostic and Statistical Manual of Mental Disorders (DSM-III-R) criteria for SCZ or poor-outcome schizoaffective disorder. Such multiplex families, display substantially higher recurrence risk of SCZ than reported in sporadic cases (8,9), and this discrepancy in recurrence risk suggests that there may be important differences in the genetic architecture between familial and sporadic SCZ cases that warrant further investigation.

One explanation of this difference is that familial SCZ cases may carry a higher burden of common SCZ risk variation as measured by a higher SCZ polygenic risk score (PRS), than ancestry matched sporadic cases. Another explanation is that the increased recurrence risk in multiplex families may be attributable to segregation of rarer, higher risk variation, identifiable through exome or whole-genome sequencing likely in combination with common risk variation. Sequencing studies suggest that rare, deleterious variation in the genome is involved in the genetic etiology of SCZ and other psychiatric disorders (15–20), but the extent to which rare variation contributes to SCZ risk in multiplex families is currently unknown. A third hypothesis, not addressed here, is that familial cases may have increased exposure to environmental risks unique to the families that may explain the higher recurrence risk in multiplex families.

Mega-analyses of SCZ genome-wide association study (GWAS) data by the Psychiatric Genomics Consortium Schizophrenia Working Group (PGC-SCZ) have identified more than 270 loci associated with SCZ (21–23). GWAS data from such studies are frequently used to construct PRS to index an individual’s common genetic variant risk for a disorder. Although current PRS currently lack power to predict SCZ in the general population, they have been shown to index meaningful differences in SCZ liability between individuals. For example, in the European PGC3-SCZ sample, the highest PRS centile has an OR of 44 (95% CI=31-63) for SCZ compared to the lowest centile of PRS, and OR of 7 (95% CI=5.8-8.3) when the top centile is compared with the remaining 99% of the individuals in the sample (23).

Common risk variation analyses in multiplex family samples smaller than ISHDSF have been performed (24–26), and we have previously used the summary statistics from the first wave of PGC-SCZ mega-analysis (21) to investigate whether the concept of the genetically influenced psychosis spectrum is supported by empirical data in multiplex SCZ families (27). Here, we extend our previous work by using PRS profiling in multiplex SCZ families, sporadic SCZ cases and population controls, all from the population of the island of Ireland, to directly test whether common SCZ risk variation in the genome may explain the increased recurrence risk of SCZ in multiplex families. Identifying the source of the increased familial recurrence risk of SCZ is important for future research into the genetic etiology of familial SCZ, and potentially for both diagnosis and treatment of SCZ with different familial backgrounds, as it will determine the relative focus on environmental exposures, as well as common and rare genetic variation in case-control and family studies of SCZ.

## Methods

### Sample Description

#### Irish Study of High-Density Schizophrenia Families (ISHDSF)

Fieldwork for the ISHDSF sample were carried out between 1987 and 1992, with probands ascertained from public psychiatric hospitals in the Republic of Ireland and Northern Ireland, with approval from local ethics committees (28). Inclusion criteria were two or more first-degree relatives meeting DSM-III-R criteria for SCZ or poor-outcome schizoaffective disorder (PO-SAD), with all four grandparents being born in Ireland or the United Kingdom. Relatives of probands suspected of having psychotic illness were interviewed by trained psychiatrists, and trained social worked interviewed other relatives. Hospital and out-patient records were obtained and abstracted in > 98% of cases with SCZ or PO-SAD diagnoses. To avoid bias and detect possible mistakes in diagnosis, independent review of all diagnostic information such as interview, family history reports, and hospital information was made blind to family assignments by two trained psychiatrists, with each psychiatrics making up to 3 best estimate DSM-III-R diagnoses, with high agreement between the two psychiatrists (weighted k= 0.94 +-0.05).

The concentric diagnostic schema of the ISHDSF shown in Table 1 and Supplementary Figure 4, includes 4 case definitions: *narrow* (SCZ, PO-SAD), *intermediate* (adding schizotypal personality, schizophreniform, and delusional disorders, atypical psychosis and good-outcome schizoaffective disorder), *broad* (adding psychotic affective illness, paranoid, avoidant and schizoid personality disorders and other disorders that significantly aggregate in relatives of probands) and *very broad* (adding any other psychiatric illness in the families). The ISHDSF sample also includes *unaffected* family members with no diagnosis of any psychiatric illness. The ISHDSF diagnostic schema is described extensively elsewhere (29).

**Table 1:**
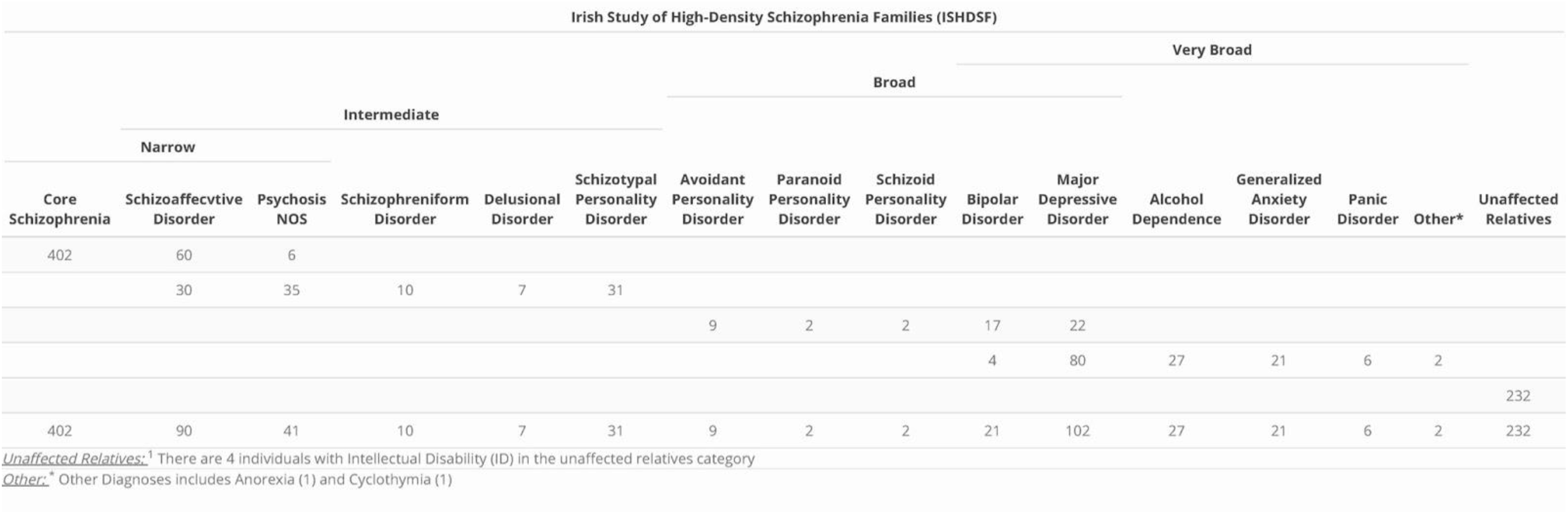
Sample description of the ISHDSF. The concentric diagnostic hierarchy of the ISHDSF contains 4 case definitions: *narrow, Intermediate, Broad* and *Very Broad*. These case definitions in the ISHDSF reflect core (narrow and intermediate) and periphery (broad and very broad) of the psychosis spectrum based on previous genetic epidemiology work referenced in the methods section. Poor-outcome schizoaffective probands are placed in the narrow category (N=60), whereas good-outcome schizoaffective probands are placed in the intermediate category (N=30).

#### Irish Schizophrenia Genomics Consortium Case/Control Sample (ISGC)

The ISGC sample was assembled for a GWAS of SCZ in Ireland. Details of recruitment, screening and quality control (QC) methods used for the ISGC sample have been previously described in detail elsewhere (30). Briefly, the case sample was recruited through community mental health service and inpatient units in the Republic of Ireland and Northern Ireland following protocols with local ethics approval. All participants were interviewed using a structured clinical interview for DSM-III-R or DSM-IV, were over 18 years of age and reported all four grandparents born either in Ireland or the United Kingdom. Cases were screened to exclude substance-induced psychotic disorder or psychosis due to a general medical condition. A subset of sporadic cases sampled by Virginia Commonwealth University (N=745) have genotypic data and FH information available (10) from completion of the family history research diagnostic criteria (FH-RDC) interview (31). This includes 233 (∼31%) FH+ cases and 512 (∼69%) FH-cases, in close concordance with the other large meta-analyses (8,9). Controls from the Irish Biobank used in ISGC were blood donors from the Irish Blood Transfusion Service recruited in the Republic of Ireland. Inclusion criteria were all four grandparents born in Ireland or the United Kingdom and no reported history of psychotic illness. Due to the relatively low lifetime prevalence of SCZ, misclassification of controls should have minimal impact on power (32).

### Genotyping and QC

Samples were genotyped using 3 different arrays (Supplementary Table 2). 830 individuals representing 237 families from the ISHDSF sample were genotyped on the Illumina 610-Quad Array. An additional 175 ISHDSF individuals from 52 families were later genotyped on the Infinium PsychArray V.1.13 Array. For the case-control sample, 1,627 sporadic cases and 1,730 controls were successfully genotyped using the Affymetrix V.6.0 Array, either at the Broad Institute or by Affymetrix. An additional 487 sporadic cases and 475 controls were later genotyped on the PsychArray along with the additional ISHDSF individuals described above. The same QC protocols were applied to all three datasets and full details are described elsewhere for ISHDSF (29) and the case-control sample (30). Exclusion criteria for samples were a call rate of <95%, more than one Mendelian error in the ISHDF sample, and difference between reported and genotypic sex. Exclusion criteria for SNPs were MAF <1%, call rate <98%, and *p*<0.0001 for deviation from Hardy-Weinberg expectation. The final ISHDSF sample included 1,005 individuals from 257 pedigrees, and the final case-control sample included 4,319 individuals (2,114 sporadic cases and 2,205 controls), whose SNP data passed all QC filters.

### Imputation

Genotypes passing QC were phased using Eagle V.2.4 (33) and phased genotypes were then imputed to the Haplotype Reference Consortium (HRC) reference panel (34) on the Michigan Imputation Server using Minimac4 (35). The HRC reference panel includes 64,975 samples from 20 different studies that are predominantly of European ancestry, making it suitable for imputation of the samples studied here. Each of the genotype sets were imputed separately and the imputed genotype probabilities were extracted and used for PRS construction and analyses. As part of the post-imputation QC, variants with MAF <1% and r^2^ score of <0.3 (36) were excluded from the analyses (Supplementary Materials and Supplementary Figures 1-3). After imputation and all QC steps, 9,298,012 SNPs in the Illumina Array, 11,080,279 SNPs in the Affymetrix Array, and 11,081,999 SNPs in the PsychArray remained for analysis. In total, 9,008,825 SNPs were shared across all three arrays and were used for PRS construction and all downstream analyses. The mean imputation quality for the SNPs used for PRS construction on each array was high (mean for all 0.96). Detailed information on imputation quality for the SNPs used for PRS construction is provided in Supplementary Materials and Supplementary Table 1.

### Construction of Polygenic Risk Scores

The ISGC and ISHDSF cohorts are part of the PGC3-SCZ GWAS. To avoid upward bias in PRS estimations, we acquired leave-N-out SCZ summary statistics from the PGC by excluding all cohorts containing any Irish subjects included in the current study. The leave-N-out GWAS summary statistics for PGC3-SCZ (N= 156,509) were first QC’d by excluding variants with MAF < 1% and imputation quality score of < 0.9, as well as removing strand ambiguous variants and insertion deletion polymorphisms. We then constructed PRS for all subjects using a Bayesian regression framework by placing a continuous shrinkage prior on SNP effect sizes using PRS-CS (37). PRS-CS uses linkage disequilibrium (LD) information from 1000 Genomes European Phase 3 European sample (38) to estimate the posterior effect sizes for each SNP. Although *p*-value thresholding method have been previously used frequently (39), PRS-CS has shown substantial improvement in predictive power compared to those methods (40).

Similar to LD Score regression (41), PRS-CS limits the SNPs for PRS construction to approximately 1.2 million variants from HapMap3. By restricting the variants to HapMap3, the partitioning provides ∼ 500 SNPs per LD block which substantially reduces memory and computational costs. The constructed PRS using PRS-CS method were normalized against the score distribution in the population control for subsequent analyses.

To show the specificity of the PRS constructed from PGC3-SCZ, an additional PRS for low density lipoprotein (LDL, N=87,048) from the ENGAGE Consortium (42) was also constructed using the same protocols described above. Genetic correlation and Mendelian Randomization studies by PGC3-SCZ show that there is no genetic correlation between SCZ and LDL, making LDL an appropriate comparison phenotype in which no inflation of SCZ PRS would be expected (43,44).

### Genomic Relationship Matrix, Principal Component and Statistical Analyses

Statistical analyses were carried out using a mixed effects logistic regression model using GMMAT package (45) in R (46). To account for the high degree of relatedness among individuals, we used *glmm*.*wald()* function, fitted by maximum likelihood using Nelder-Mead optimization. We modeled the family structure as a random effect with genetic relationship matrix (GRM) calculated using LDAK (47). In order to account for batch effects due to genotyping carried out on different arrays or at different sites, we included platform and site as covariates in the model (Supplementary Materials). Principal component analysis (PCA) of the full sample (Supplementary Materials) shows that all individuals in the sample are of European ancestry (Supplementary Figures 5-7). To account for fine-scale structure within the Irish population (Supplementary Figure 8), the top 10 principal components (PC) were also included as covariates in the analyses. The final regression models included GRM as a random effect covariate, with the top 10 PCs, genotyping platform, site, and sex as fixed effect covariates. The final results were adjusted for multiple testing using the Holm method in R.

## Results

The mean PRS across the diagnostic categories for SCZ are displayed in Figure 1. No significant differences in LDL PRS were observed between any of the diagnostic categories compared to population controls (Supplementary Figure 9), indicating the specificity of PGC3-SCZ PRS in this study. PGC3-SCZ PRS results show that the *Narrow* spectrum category in the families, which includes familial cases of SCZ, had the highest mean PRS (Z=1.13, SE=0.09) followed by sporadic cases (Z=1.06, SE=0.09), *intermediate* spectrum familial cases (Z=0.81, SE=0.10), *broad* familial spectrum cases (Z=0.67, SE=0.11), *very-broad* spectrum cases (Z=0.53, SE=0.098), *unaffected* family members (Z=0.36, SE=0.10) and population controls (Z=0.004, SE=0.07).

**Figure 1:**
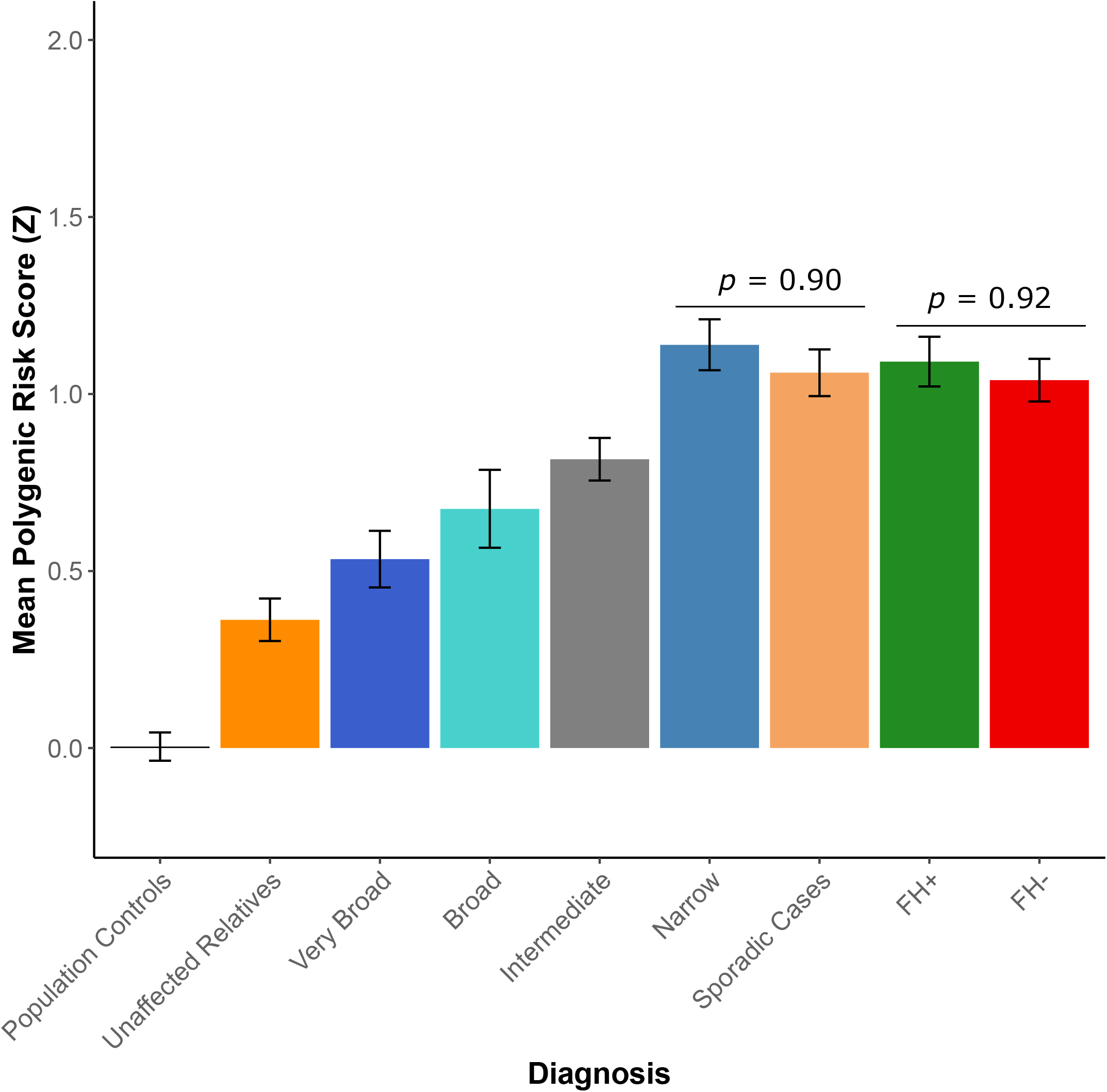
Mean PGC3-SCZ PRS for each of the diagnostic categories in the ISHDSF sample, sporadic SCZ cases and ancestry-matched population controls. A subset of sporadic cases with FH information are further divided into FH+ (green bar) and FH- (red bar) categories. Unaffected relatives (dark orange bar) are distinct from population controls (black bar) as they represent unaffected individuals in the families. Familial SCZ cases are in the narrow category as described in the methods section. Error bars represent the standard error of the observed mean. X-axis shows each of the diagnostic categories. Y-axis shows the mean normalized Z-score for PGC3-SCZ.

### No significant difference between familial and sporadic cases of SCZ

We observe no significant difference in PRS between familial SCZ cases and all sporadic SCZ cases, (*p* = 0.90), nor between familial SCZ cases and either FH+ (*p* = 0.88) or FH- (*p* = 0.82) sporadic SCZ cases. These results suggests that an increased burden of common SCZ risk variation is unlikely to account for the higher recurrence risk of SCZ in multiplex families (Figure 1). Additionally, we show that there is no significant difference in SCZ PRS between FH+ and FH-sporadic SCZ cases (*p* = 0.92), suggesting that the inclusion of all sporadic cases in the comparison is unlikely to cause an upward bias in the mean PRS for the full cohort of sporadic cases, and further supporting the hypothesis that increased PRS is unlikely to account for FH of SCZ in the cohort studied here.

### All family members carry a high burden of common SCZ risk variants

Familial and sporadic narrow SCZ cases show a significantly higher mean PRS compared to all other diagnostic categories in the ISHDSF sample and ancestry-matched population controls (Figures 1 and 2, Supplementary Table 3), underlining the important role of common risk variation in the genetic architecture of both familial and sporadic SCZ cases. All other ISHDSF diagnostic categories also show a significantly higher mean SCZ PRS than observed in the population controls (Figures). PRS comparison within the ISHDSF sample (Supplementary Table 4) shows no significant difference between mean PRS for *intermediate* and *broad* categories, indicating that individuals in both categories have a similar burden of common SCZ risk variants despite the presence of a range of diagnoses on the psychosis spectrum such as atypical psychosis and delusional disorder in the *intermediate* category, and disorders such as major depressive disorder with psychotic features, and bipolar disorder in the *broad* category. We observed no significant difference between the *broad* category and the *very-broad* category, which includes any other psychiatric disorder in the ISHDSF sample. The mean SCZ PRS in the *very broad* category is not significantly different from the *unaffected* members of the families, indicating a similar burden of common SCZ risk variation in these two distinct diagnostic categories. Finally, we observe a significantly higher PRS in *unaffected* family members compared to the population controls (*P* = 4.13 × 10^−3^), indicating a high baseline risk for SCZ in all members of multiplex families compared to population controls, regardless of their diagnostic status. This observation is consistent with SCZ transmission through some unaffected family members observed in the ISHDSF and other family samples

**Figure 2:**
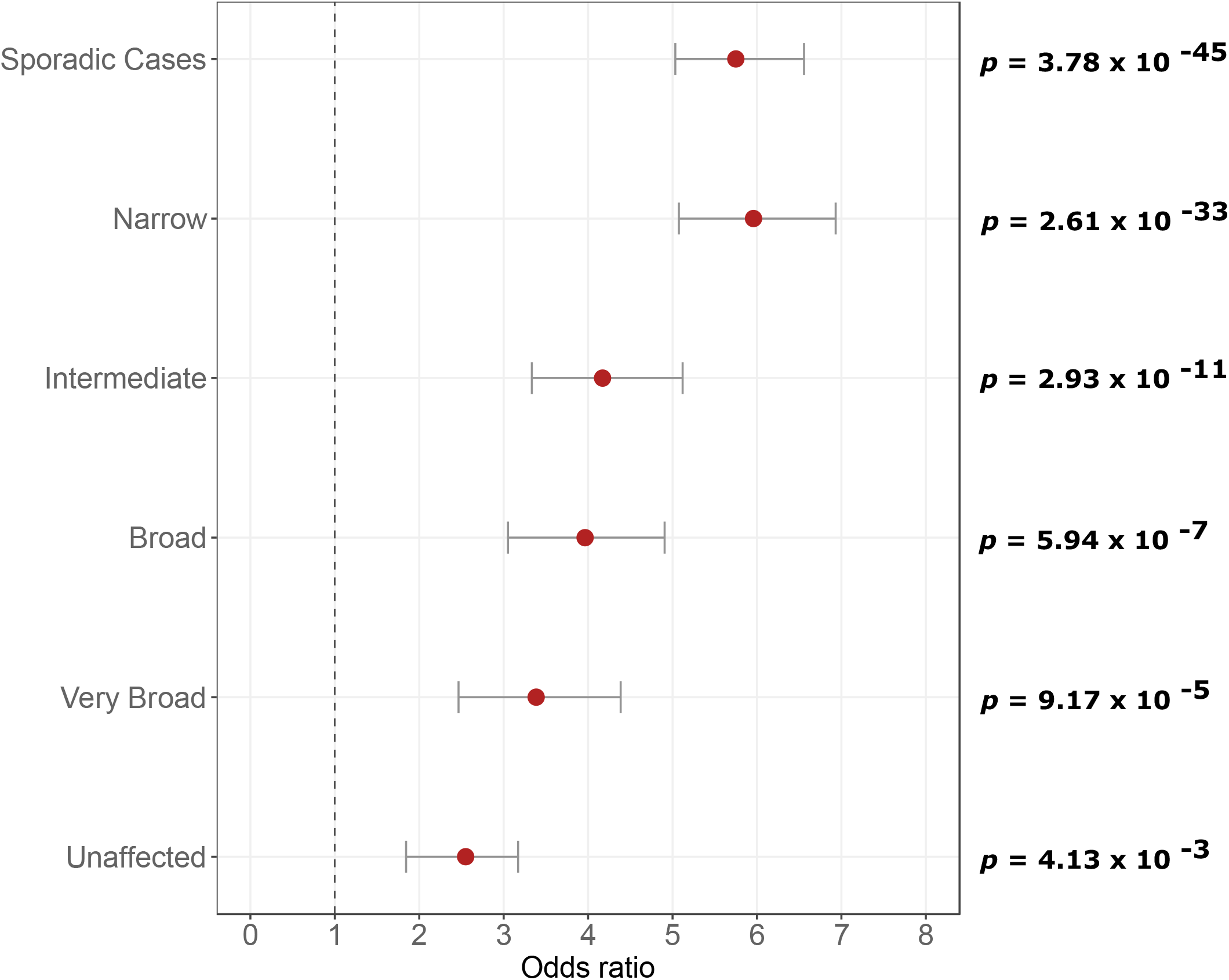
Comparison of PRS between ISHDSF diagnostic categories and sporadic cases versus population controls. All analyses follow the hypothesis that ISHDSF members and sporadic cases have a higher PRS compared to population controls. All PRS have been normalized using Z-score standardization prior to obtaining odds ratios. The plots show odds ratios (OR, filled circles) with 95% confidence intervals (CI) for each category compared to population controls. *p-*values after multiple testing correction are provided on the right side of the Y-axis. X-axis represents the odds ratios. Left side of Y-axis represents each of the categories used for comparison versus population controls.

## Discussion

Multiplex SCZ families represent the upper bounds of the distribution of recurrence risk for SCZ, and this study aimed to investigate the source of this increased recurrence risk. Since sporadic cases are considered to be the norm for most complex diseases including SCZ (48), this makes sporadic SCZ cases a good comparison group to assess whether elevated PRS can account for the increase in recurrence risk in familial cases. We observed that familial SCZ cases do not have a significantly increased PRS compared to sporadic SCZ cases in our modestly sized sample. We further show that this observation holds true regardless of the FH status of sporadic cases. Therefore, our finding provides empirical evidence that increased recurrence risk of SCZ in the ISHDSF sample is unlikely to be attributable to an increased burden of common SCZ risk variation as identified from genome-wide association studies. Therefore, the hypothesis that high familial recurrence risk of SCZ in multiplex families may be attributable to excess rare variation in the genome specific to SCZ, warrants further investigation. Furthermore, these results validate the concept of a genetically influenced psychosis spectrum in multiplex SCZ families as shown by a continuous increase of common SCZ risk variation burden across all members of the ISHDSF, from *unaffected* family members, to *narrow* category in the ISHDSF sample. This analysis reveals potentially important differences in the genetic architecture of familial SCZ cases compared to familial bipolar disorder (BIP) cases. Analysis conducted by Andlauer et al (25) on BIP multiplex families have shown that unlike familial SCZ cases studied here, familial BIP cases have a significantly higher BIP PRS compared to ancestry matched, sporadic cases. These results, in addition to sparse evidence for the involvement of rare risk variation in the genetic architecture of BIP, demonstrates the importance of common risk variation in familial BIP, whereas whole exome sequencing studies of SCZ in both family and case-control samples have demonstrated that rare variation is likely to play an important role in SCZ risk (49–52).

Although sequencing studies are only now reaching sample sizes sufficiently powered to detect individually associated rare variation and rare variant enriched genes associated with SCZ (52), earlier sequencing and rare variation studies observe consistent enrichment of rare variation in certain gene-sets and functional categories related to SCZ (49). In addition, SNP signals from PGC3-SCZ GWAS are shown to be highly enriched in noncoding functional sequences in the genome (23), further underscoring the importance of conducting large scale whole-genome sequencing to identify rare variation in non-coding regions of the genome linked to SCZ. Results from the 1000 Genomes Project demonstrates that rare functional variation is frequent in the genome (53) and shows strong population specificity (54). For example, using GWAS probe intensity data in the Irish case-control sample used in this study, we have previously detected a rare, novel 149kb duplication overlapping the protein activated kinase 7 (*PAK7*) gene only found in the Irish population (55). This duplication is associated with SCZ in the ISGC (*p* = 0.007), and a replication sample of Irish and UK case-controls with 22 carriers in 11,707 cases and 10 carriers in 21,204 controls (*p* = 0.0004, OR=11.3). This duplication in *PAK7* gene is in strong LD with local haplotypes (*p* = 2.5 × 10^−21^), indicating a single ancestral event and inheritance identical by descent in carriers.

We note that the liability that is captured by PRS constructed from PGC3-SCZ is currently insufficient for predicting a diagnosis of SCZ (AUC=0.71) (23), meaning that PRS alone cannot be used as a diagnostic tool. The results of our study further suggest that current PRS alone is unlikely to be predictive of SCZ recurrence risk in the families of index probands. To address both of these predictive limitations of SCZ PRS, additional components of genetic risk must be identified and included in order to improve both identification of future cases and recurrence risk prediction in the relatives of probands.

The results presented in this study should be interpreted in the context of some limitations. First, current PGC3-SCZ PRS accounts for ∼2.6% of the total variance in SCZ liability (23), and genetic risks from rare and structural variation are not represented in the PRS. As a result, some known genetic risk factors for SCZ such as the 22-q11 deletion (56) are not included in PRS construction, and such genetic risk factors are best measured through direct assessment of structural variation or whole genome sequencing studies. Despite these limitations, PRS provide the most reliable measurement of common risk variation in the genome and are suitable for indexing an individual’s risk for SCZ in this study. Second, the various diagnostic categories in the ISHDSF sample contain different number of subjects (28). For example, the lower number of individuals satisfying *broad* and *very broad* diagnostic schema in the families, means that the power of analysis in those subgroups is lower. However, the *narrow* category which includes familial SCZ cases in the ISHDSF sample, has the highest number of individuals across all the diagnostic categories in the ISHDSF, making the sample suitable for the main hypothesis being tested in this study. Third, FH information is only available for a subset of sporadic cases as described in the methods.

However, the ratio of FH+ (∼31%) and FH- (∼69%) sporadic cases studied here is in close agreement with FH data from large meta-analyses samples (8,9), suggesting the subset of sporadic FH+ cases available are representative. Fourth, this analysis did not assess the common risk variant burden of each family separately, and the degree to which common risk variation may impact each family could vary between different families. Fifth, since the environmental factors unique to the families have also not been systematically assessed here, integrating rare genetic variation from whole sequencing studies with environmental influences in future analyses could further elucidate the role of rare variation and environmental influences on the recurrence risk of SCZ in multiplex families. Finally, as more samples from under-represented populations are collected, it is essential to replicate and show the generalizability of these findings in more diverse populations.

In conclusion, in this study, we show that differences in common risk variation as indexed by current PRS, is unlikely to account for the increased recurrence risk of SCZ in our cohort of multiplex SCZ families and ancestry matched sporadic cases. Therefore, our results suggest that both common and rare SCZ risk variation needs to be indexed to potentially improve diagnostic and familial recurrence prediction of SCZ.

## Data Availability

GWAS summary statistics for PGC3-SCZ GWAS is publicly available ON PGC website https://www.med.unc.edu/pgc/download-results/
Leave-N-out PGC3-SCZ GWAS summary statistics excluding the Irish samples can be obtained by contacting the corresponding author
GWAS summary statistics for LDL is publicly available at ENGAGE Consortium website http://diagram-consortium.org/2015_ENGAGE_1KG/.
We made use of various freely available software tools in this study:
PRS-CS: https://github.com/getian107/PRScs
PLINK: https://www.cog-genomics.org/plink/2.0/
GMMAT: https://github.com/hanchenphd/GMMAT
LDAK: http://dougspeed.com/ldak/
The custom scripts used in this study will be made publicly available upon publication.

## Data Availability

GWAS summary statistics for PGC3-SCZ GWAS is publicly available on the PGC website https://www.med.unc.edu/pgc/download-results/

Leave-N-out GWAS summary statistics for PGC3-SCZ GWAS was acquired from the PGC by following the appropriate guidelines and will be shared upon reasonable request.

GWAS summary statistics for LDL is publicly available on the ENGAGE Consortium website http://diagram-consortium.org/2015_ENGAGE_1KG/

We made use of various freely available software tools in this study:

PRS-CS: https://github.com/getian107/PRScs

PLINK: https://www.cog-genomics.org/plink/2.0/

GMMAT: https://github.com/hanchenphd/GMMAT

LDAK: http://dougspeed.com/ldak/

The custom scripts used in this study are available upon request.

## Funding and Acknowledgements

MA, BV, S-AB, KSK, BTW and BR were supported by R01-MH114593 (to Dr. Riley). Production of GWAS data for sporadic cases and controls was supported by R01-MH083094 (to Dr. Riley) and Wellcome Trust Case Control Consortium 2 project (085475/B/08/Z and 085475/Z/08/Z), and the Wellcome Trust (072894/Z/03/Z, 090532/Z/09/Z and 075491/Z/04/B), and NIMH grant MH 41953 and Science Foundation Ireland (08/IN.1/B1916). Production of GWAS data for multiplex families was supported by R01-MH062276 and R01-MH068881 (to Dr. Riley). AEG was supported by T32-MH020030. T-HN was supported by NARSAD Young Investigator Grant 28599.

## Membership of the Irish Schizophrenia Genomics Consortium (ISGC)

Brien P Riley^1^, Derek W Morris^2^, Colm T O’Dushlaine^3^, Paul Cormican^4^, Elaine M Kenny^3^, Brandon Wormley^1^, Gary Donohoe^2^, Emma Quinn^3^, Roisin Judge^3^, Kim Coleman^3^, Daniela Tropea^3^, Siobhan Roche^5^, Liz Cummings^3^, Eric Kelleher^3^, Patrick McKeon^5^, Ted Dinan^6^, Colm McDonald^2^, Kieran C Murphy^7^, Eadbhard O’Callaghan^8^, Francis A O’Neill^9^, John L Waddington^10^, Ken S Kendler^1^, Michael Gill^3^, Aiden Corvin^3^

1 Depts of Psychiatry and Human Genetics, Virginia Institute of Psychiatric and Behavioral Genetics, Virginia Commonwealth University, Richmond, VA, USA;

2 Centre for Neuroimaging and Cognitive Genomics (NICOG), School of Psychology, National University of Ireland Galway, Ireland; School of Natural Sciences, National University of Ireland Galway, Ireland;

^3^ Neuropsychiatric Genetics Research Group, Institute of Molecular Medicine, Trinity College Dublin, Dublin, Ireland;

^4^ Animal and Grassland Research and Innovation Centre, Teagasc, Grange, Dunsany, County Meath, Ireland;

^5^ St Patrick’s University Hospital, James St., Dublin, Ireland;

^6^ Department of Psychiatry, University College Cork, Cork, Ireland;

^7^ Department of Psychiatry, Royal College of Surgeons in Ireland, Dublin, Ireland;

^8^ DETECT Early Psychosis Service, Blackrock, Co. Dublin, Ireland;

^9^ Centre for Public Health, Institute of Clinical Sciences, Queen’s University Belfast, Belfast, UK;

^10^ Molecular and Cellular Therapeutics, Royal College of Surgeons in Ireland, Dublin, Ireland;

